# Socioeconomic Vulnerability, Social Protection, and HIV Outcomes Among Pregnant and Lactating Women in Zimbabwe

**DOI:** 10.64898/2026.03.15.26348437

**Authors:** Mollie Hudson, Rutendo Mukondwa, Cesar Aviles-Guaman, Amrita Ayer, Kudakwashe Takarinda, Tatenda Makoni, Sinokuthaba Mukungwa, Waraidzo Mukuwapasi, Karen Webb, Priya B. Shete

## Abstract

**Background:** Pregnant and lactating women with HIV are particularly vulnerable to the intersecting effects of HIV and poverty. In Zimbabwe, socioeconomic vulnerability among young women increases the risk of HIV acquisition and compromises treatment outcomes. Addressing poverty is therefore essential to improving maternal and child health outcomes in this population. This retrospective cross-sectional study examined the relationship between socioeconomic vulnerability and HIV-related health outcomes among pregnant and lactating women in Zimbabwe.

**Methods:** We surveyed 600 pregnant and lactating women living with HIV receiving care at community health centers across Zimbabwe. The survey captured sociodemographic and self-reported clinical data. Using generalized estimating equations, we assessed associations between viral load non-suppression, antiretroviral (ARV) treatment interruption, delayed presentation to antenatal care, and socioeconomic covariates including poverty level and participation in social protection programs.

**Results:** Among participants, 74.5% (n=447) were lactating and 25.5% (n=153) were pregnant. Over half (54.2%, n=325) reported at least one unmet social need; 47.2% (n=238) experienced food insecurity, 26.2% (n=157) lacked transportation funds, and 23.7% (n=142) lacked money for user fees or related health costs. Nearly half (49.5%, n=297) reported using negative financial coping strategies such as taking loans, selling assets, or withdrawing savings. Intimate partner violence was significantly associated with viral load non-suppression (RR 2.4, 95% CI 1.5–4.0, p<0.005). Receipt of social protection benefits was associated with ARV treatment interruption (RR 2.3, 95% CI 1.3–4.2, p=0.006), while dissaving was associated with delayed antenatal care presentation (RR 1.8, 95% CI 1.4–2.3, p<0.05).

**Conclusions:** Unmet social needs, poverty, and adverse financial coping strategies were associated with suboptimal HIV outcomes among pregnant and lactating women in Zimbabwe. Although social protection interventions aim to mitigate these vulnerabilities, their complex effects warrant further investigation to optimize design and implementation.

## INTRODUCTION

Human Immunodeficiency Virus (HIV) remains a leading cause of death in sub-Saharan Africa. ^1^In 2021, of the 650,000 AIDS-related deaths reported globally, 280,000 occurred in sub-Saharan Africa.^2^ Pregnant women and women of reproductive age are disproportionately affected by the HIV epidemic. For instance, in South Africa, HIV prevalence among women aged 15–29 years is approximately 22.6%, compared to 11.5% among men aged 15–49.^3^ In Lesotho, women aged 20–24 years have nearly four times the HIV prevalence of men in the same age group.^4^ Of the estimated 1.4 million annual pregnancies among women living with HIV, 90% occur in sub-Saharan Africa.^5^ A 2022 study in Malawi found that 30% of women admitted for antenatal care at a primary hospital were HIV-positive,^6^ while in South Africa, one in three pregnant women is living with HIV.^3^ Despite universal access to antiretroviral (ARV) therapy, HIV-positive pregnant women remain more likely to experience intrauterine fetal death or low birth weight deliveries compared to HIV-negative counterparts.^7^

In Zimbabwe, HIV prevalence is similarly higher among women than men—15.4% versus 11.3%, respectively.^8^ Among individuals aged 15–29 years, women have approximately six times the HIV incidence of men in the same age cohort.^9^ Socioeconomic determinants of health, including poverty, early marriage, and limited access to education, have been shown to drive HIV risk in this population. A 2015 study found that lower socioeconomic status was associated with earlier marriage and higher engagement in risky sexual behaviors, as well as with increased food insecurity and HIV infection.^10^ Poverty also poses a significant barrier to antenatal care engagement, contributing to higher rates of mother-to-child transmission of HIV.^11^ Pregnant and lactating women are particularly susceptible to the effects of poverty; food insecurity during pregnancy is linked to adverse physical and mental health outcomes,^12^ and the nutritional demands of lactation further heighten vulnerability to food insecurity.^13^ Together, these findings underscore the critical importance of addressing poverty to improve maternal health outcomes among women living with HIV.

Although previous research has described the relationship between HIV and poverty and the heightened vulnerability of pregnant and lactating women,^10–12^ there remains limited evidence quantifying how specific dimensions of socioeconomic vulnerability—or unmet social needs—affect HIV-related outcomes in this group. Moreover, little is known about the reach, inclusivity, and coverage of interventions designed to mitigate socioeconomic vulnerability, commonly referred to as social protection interventions, among pregnant and lactating people. This study aimed to examine the association between multiple measures of socioeconomic vulnerability and key clinical outcomes among pregnant and lactating women living with HIV in Zimbabwe—a country with high levels of poverty and food insecurity^14^ and a generalized HIV epidemic. Additionally, we sought to describe the extent of social protection program coverage in this population and explore how these interventions may influence HIV treatment outcomes and engagement in antenatal care.

## METHODS

### Study Design

We conducted a retrospective cross-sectional study using data from a Client Satisfaction Survey (CSS) implemented between February 8 and July 27, 2023, by the Organization for Public Health Interventions and Development (OPHID). The CSS is a routine quarterly survey used by OPHID and partners to assess the quality of HIV care at participating health facilities across 15 districts in Zimbabwe. The survey is administered in English, Shona, or Ndebele by Community HIV and AIDS Support Agents (CHASAs), who are staff members of the Zimbabwe National Network of People Living with HIV (ZNNP+). ZNNP+ is an umbrella organization that coordinates HIV support groups across the country and collaborates with OPHID on multiple programmatic activities.^15^ Surveys typically take 30–45 minutes to complete. For this study, we appended additional questions to the standard CSS tool to collect data on self-reported clinical outcomes, unmet social needs (e.g., food insecurity, transportation or healthcare costs), and prior receipt of social protection interventions.

### Study Setting and Population

Participants were systematically sampled from 317 OPHID-supported health centers across 15 districts in Zimbabwe using stratified random sampling to ensure representation by geographic and facility characteristics. Health facilities were able to Eligible participants were adults aged ≥18 years, living with HIV, receiving HIV care at an OPHID-supported facility, and either pregnant or lactating at the time of data collection. Sample size was determined by feasibility within routine CSS operations.

### HIV Treatment Outcomes

Two self-reported HIV treatment outcomes were assessed: (1) VIRAL LOAD NON-SUPPRESSION, and (2) ARV TREATMENT INTERRUPTION. Viral load non-suppression was defined dichotomously as ≥1,000 copies/mL versus <1,000 copies/mL, consistent with national laboratory thresholds in Zimbabwe, where assays cannot detect viral loads below 1,000 copies/mL.^16^ Individuals with viral loads <1,000 copies/mL were categorized as having controlled viral suppression. ARV treatment interruption was defined as any self-reported discontinuation of ARVs for any duration (yes/no). Both outcomes represent routinely monitored indicators within HIV primary care at OPHID-supported facilities.

### Antenatal Care Outcomes

Engagement in antenatal care (ANC) was evaluated by self-reported timing of first ANC visit. Women initiating ANC before 12 weeks of gestation were considered to have *timely* presentation, while those presenting after 12 weeks were classified as having *delayed presentation*.^17^

### Covariates

We collected data on demographic, social, and clinical characteristics, including household size, disability status, travel time to clinic, healthcare-related costs, and DISSAVING (a negative financial coping strategy encompassing selling assets, taking loans, withdrawing savings, unenrolling a child from school, or reducing household food consumption). Additional variables included receipt and type of social protection, unmet social needs (e.g., food insecurity, lack of transportation funds, lack of healthcare funds, or other forms of socioeconomic vulnerability), multidimensional poverty index (MPI) quartile based on ten indicators across health, education, and living standards,^18^ intimate partner violence, gender-based violence, and number of months of ARVs dispensed. Covariate definitions are detailed in APPENDIX A.

### Statistical Analysis

Descriptive statistics summarized participants’ demographic, social, and clinical characteristics (**Table 1; Supplementary Table 1**). We stratified participants by receipt of social protection (**Supplementary Table 2**) and compared groups across covariates. Generalized estimating equation (GEE) models with district-level clustering were used to assess associations between socioeconomic factors and health outcomes, accounting for district-level variation in socioeconomic risk and social protection program coverage.^19^

**Table 1.**
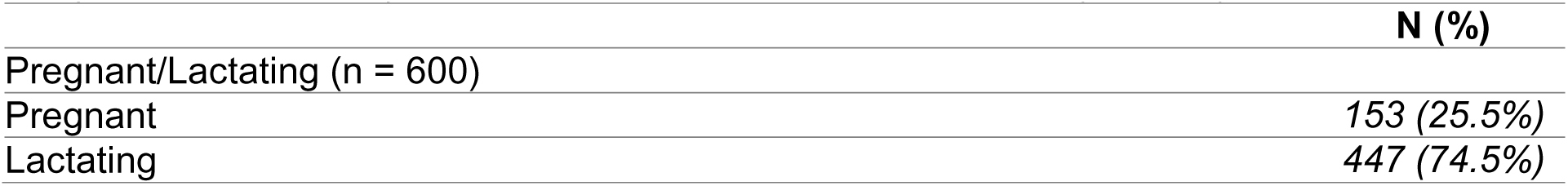
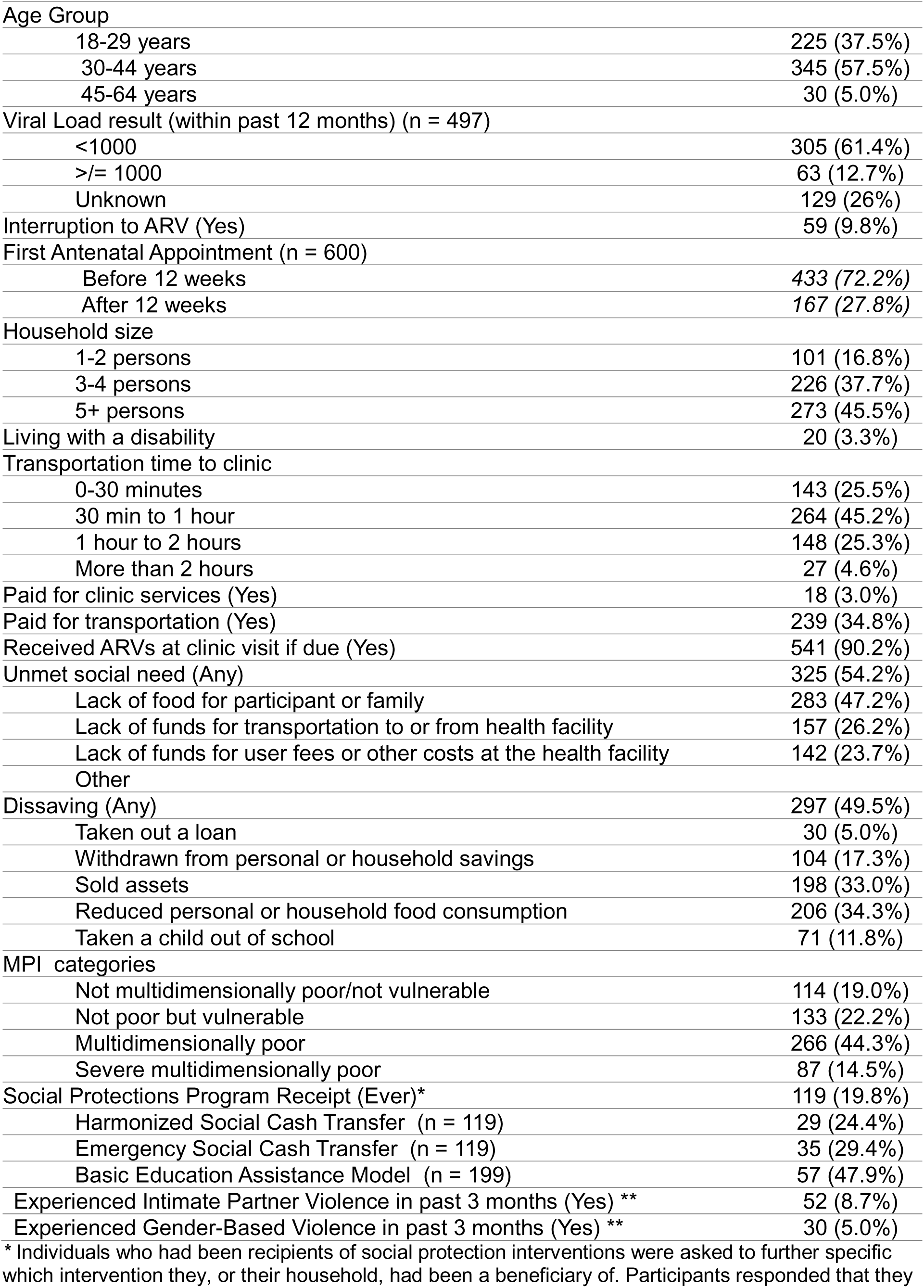
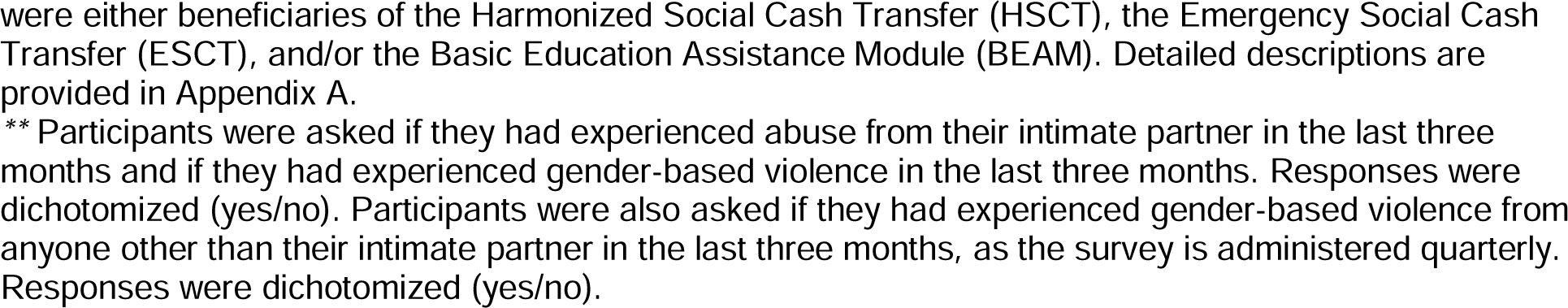
Demographics and socioeconomic characteristics of pregnant and lactating women living with and receiving care for HIV in 15 districts of Zimbabwe (N = 600)

Independent variables included household size, disability, travel time to clinic, dissaving, costs incurred with care, MPI, social protection receipt, and exposure to intimate partner or gender-based violence. Dependent variables were (Model 1) HIV viral load non-suppression, (Model 2) ARV treatment interruption, and (Model 3) delayed ANC presentation. For Model 1, ARV interruption was also included as a covariate given its expected association with viral load outcomes.^20^ Both unadjusted and adjusted models were fitted. Multivariate Poisson regression with a log link function was used to estimate risk ratios (RR) and 95% confidence intervals (CI), with p<0.05 considered statistically significant. Analyses were performed using STATA BE VERSION 17.0.^21^

### Ethical Considerations

The study was approved by the Institutional Review Boards of the University of California, San Francisco (IRB # 21-34325), and the Medical Research Council of Zimbabwe (protocol # MRCZ/A/2888). Written informed consent was waived as the primary purpose of the CSS is quality-of-care monitoring. Only de-identified data were analyzed.

## RESULTS

### Descriptive statistics

A total of 600 participants were surveyed across 15 districts in Zimbabwe (**Table 1; Supplementary Table 1**). Most respondents (57.5%, n=345) were aged 30–44 years, and 37.5% (n=224) were aged 18–29 years. Participants were relatively evenly distributed across districts, except for Bulawayo, which contributed 2.2% (n=13) of the sample. The majority (74.5%, n=447) were lactating at the time of the survey, while 25.5% (n=153) were pregnant. Nearly three-quarters (72.2%, n=433) reported attending their first antenatal appointment within 12 weeks of pregnancy.

Almost half of participants (45.2%, n=264) reported traveling 30 minutes to one hour to reach a health facility. Of the 600 respondents, 90% (n=541) had received a viral load test result within the past 12 months. Among these, 61.4% (n=305) reported viral load suppression (<1,000 copies/mL), while 26% (n=129) did not know their most recent viral load result.

More than half of participants (54.2%, n=325) reported at least one form of socioeconomic vulnerability that affected their ability to adhere to ART or attend clinic visits. The most common challenge was food insecurity, reported by 47.2% (n=238) of respondents. One in four participants reported lacking funds for transportation to or from a health facility (26.2%, n=157) or for user fees and other healthcare-related expenses (23.7%, n=142). Nearly half (49.5%, n=297) reported engaging in dissaving, defined as taking loans, selling assets, or withdrawing savings to cope with financial strain.

When stratified by receipt of social protection, sociodemographic and clinical characteristics were broadly similar between groups (**Supplementary Table 2**). However, recipients of social protection were less likely to report viral load suppression compared with non-recipients (70.6%, n=48 vs. 85.7%, n=257; *p*=<0.05). The prevalence of unmet social needs was similar between the two groups (53.3% vs. 53.6%; *p*=0.6). Recipients of social protection interventions reported slightly higher rates of dissaving (55.5%, n=66 vs. 48.0%, n=231), although this difference was not statistically significant (*p*=0.1).

### HIV viral load non-suppression

Results of the GEE models examining factors associated with HIV viral load non-suppression are shown in **Table 2**. In the unadjusted model, several socioeconomic and psychosocial characteristics were significantly associated with viral load non-suppression. These included living with a disability (RR 2.3; 95% CI 1.2–4.4; *p*=<0.05), receipt of social protection benefits (RR 2.1; 95% CI 1.3–3.4; *p*=<0.05), and recent experience of intimate partner violence (RR 2.6; 95% CI 1.3–5.6; *p*=<0.05). After adjustment for potential confounders, only intimate partner violence remained significantly associated with viral load non-suppression (adjusted RR 2.4; 95% CI 1.5–4.0; *p*<0.05). Household size, travel time to clinic, costs incurred with care, and ART interruption were not significantly associated with viral load non-suppression in either the unadjusted or adjusted models.

**Table 2.** Association of patient characteristics with HIV viral load non-suppression (n=361), controlling for age.

|  | Unadjusted<br>RR (95% CI) | p-value | Adjusted RR<br>(95% CI) | p-value |
| --- | --- | --- | --- | --- |
| Household size |  |  |  |  |
| 3-4 | 1 (.42, 2.4) | 0.9 | 0.82 (.32, 2.1) | 0.7 |
| 5+ | 1.2 (.5, 2.6) | 0.7 | 1.0 (0.5, 2.3) | 0.9 |
| Living with a disability | 2.3 (1.2, 4.4) | 0.01 | 1.8 (.9, 3.5) | 0.1 |
| Transportation time to<br>clinic |  |  |  |  |
| 0-30 min (Ref) | -- | - | -- | - |
| 30 min to 1 hour | 1.7 (.8, 3.6) | 0.1 | 1.6 (.8, 3.0) | 0.2 |
| 1 hour to 2 hours | 1.1 (.4, 2.7) | 0.9 | 1 (.3, 2.9) | 1.0 |
| More than 2 hours | 1.8 (.5,6.4) | 0.4 | 1.7 (.7, 4.2) | 0.3 |
| Costs incurred with care<br>(transportation or clinic costs) | 1.1 (.7, 1.8) | 0.7 | 1.1 (0.7, 2.0) | 0.6 |
| Dissaving | 1.0 (.5, 2.1) | 1 | 1 (.5, 1.8) | 0.9 |
| Unmet social need | 0.7 (.4, 1.4) | 0.4 | .7 (.4, 1.2) | 0.7 |
| Multidimensional poverty index |  |  |  |  |
| Not<br>multidimensionally<br>poor/not vulnerable (Ref.) | --. | -- | -- | -- |
| Not<br>multidimensionally<br>poor/vulnerable | 0.8 (.3, 1.7) | 0.5 | 0.9 (.4, 1.9) | 0.8 |
| Multidimensionally<br>poor/severely poor | 1.3 (.6, 2.8) | 0.5 | 1.3 (.7, 2.7) | 0.4 |
| Social Protections<br>Receipt | 2.1 (1.3, 3.4) | <.05<br>(0.004) | 1.1 (.3, 3.5) | 0.9 |
| Interruption to ARV | 0.7 (.2, 1.9) | 0.5 | .48 (.1, 1.9) | 0.3 |
|  | Unadjusted<br>RR (95% CI) | p-value | Adjusted RR<br>(95% CI) | p-value |
| Gender based violence | 1.2 (.4, 3.5) | 0.7 | .7 (.3, 1.5) | 0.4 |
| Intimate partner violence | 2.6 (1.3, 5.6) | <.05 | 2.4 (1.5, 4) | <.05 |

### ARV treatment interruption

Model 2 (**Table 3**) estimated associations between socioeconomic characteristics and ARV treatment interruption. In the unadjusted model, having more than five individuals in the household was significantly associated with ARV interruption (RR 2.6; 95% CI 1.1–5.8; *p*=<0.05). In the adjusted model, receipt of social protection emerged as the only significant predictor of ARV interruption (adjusted RR 2.3; 95% CI 1.3–4.2; *p*=<.05).

**Table 3.** Association between patient sociodemographic and clinical characteristics with antiretroviral (ARV) interruption (n=582)

|  | Unadjusted RR<br>(95% CI) | p-value | Adjusted RR<br>(95% CI) | p-value |
| --- | --- | --- | --- | --- |
| Household size |  |  |  |  |
| 3-4 | 1.8 (0.9, 3.8) | 0.12 | 1.7 (.7, 4.1) | 0.2 |
| 5+ | 2.6 (1.1, 5.8) | <.05<br>(0.03) | 2.3 (1, 5.7) | 0.1 |
| Living with a disability | 1 (.4, 2.4) | 1 | .79 (.4, 1.7) | 0.5 |
| Transportation time to<br>clinic |  |  |  |  |
| 0-30 min (Ref) | -- | -- | -- | -- |
| 30 min to 1 hour | 1 (.5, 2.1) | 0.9 | .9 (.4, 1.9) | 0.8 |
| 1 hour to 2 hours | 1.1 (.5, 2.4) | 0.8 | .9 (.4, 2.0) | 0.8 |
| More than 2 hours | 2.4 (.7, 8.4) | 0.2 | 2.1 (.5, 8.9) | 0.3 |
| Costs incurred with<br>care (transportation or clinic<br>costs) | 1.1 (.7, 1.8) | 0.6 | 1.1 (0.6, 1.8) | 0.9 |
| Dissaving | 1 (.7, 1.5) | 0.9 | .9 (.6, 1.3) | 0.6 |
| Unmet social need | 1.1 (.7, 1.7) | 0.8 | 1.1 (.6, 1.8) | 0.8 |
| MPI 3 Categories |  |  |  |  |
| Not multidimensionally poor/not vulnerable (Ref.) | -- | -- | -- | -- |
| Not multidimensionally poor/vulnerable | 1 (0.5, 2) | 0.9 | 1 (.5, 1.9) | 1.0 |
| Multidimensionally poor/severely poor | 1.1 (0.6, 2.1) | 0.8 | 1.2 (.7, 2.0) | 0.8 |
| Social Protections Receipt | 1.7 (0.9, 3) | 0.1 | 2.3 (1.3, 4.2) | <0.05 |
| Gender based violence | 1.3 (0.6, 3.1) | 0.5 | 1 (.5, 2.1) | 1 |
| Intimate partner violence | 1.4 (0.5, 3.9) | 0.6 | 1.6 (.5, 4.7) | 0.4 |

### Delayed presentation to antenatal care

Model 3 (**Table 4**) examined factors associated with delayed presentation to antenatal care. In both unadjusted and adjusted models, dissaving was significantly associated with delayed presentation (unadjusted RR 1.7; 95% CI 1.4–2.3; *p*<0.05; adjusted RR 1.8; 95% CI 1.4–2.3; *p*<0.05). Similarly, intimate partner violence was associated with delayed antenatal presentation in both models (unadjusted RR 1.9; 95% CI 1.4–2.5; *p*<0.05; adjusted RR 1.6; 95% CI 1.0–2.4; *p*=<0.05).

**Table 4.** Association between patient characteristics and delayed presentation to antenatal care (> 12 weeks) (n=582)

|  | Unadjusted RR (95% CI) | p-value | Adjusted RR (95% CI) | p-value |
| --- | --- | --- | --- | --- |
| Household size |  |  |  |  |
| 3-4 | 0.8 (.6, 1.2) | 0.2 | 0.8 (.4, 1.6) | 0.5 |
| 5+ | 1 (.8, 1.4) | 0.8 | 1.0 (.6, 1.1) | 0.8 |
| Living with a disability | 1.3 (.9, 1.9) | 0.2 | 1.2 (.9, 1.7) | 0.3 |
| Transportation time to clinic |  |  |  |  |
| 0-30 min (Ref) | -- | -- | -- | -- |
| 30 min to 1 hour | 0.8 (.5, 1.1) | 0.2 | 0.7 (.5, 1.0) | 0.1 |
| 1 hour to 2 hours | 1 (.8, 1.2) | 0.8 | .9 (.7, 1.1) | 0.2 |
| More than 2 hours | 0.8 (.5, 1.3) | 0.4 | .8 (.5, 1.4) | 0.4 |
| Costs incurred with care (transportation or clinic costs) | 1.2 (.9, 1.5) | 0.2 | 1.1 (0.6, 1.8) | 0.9 |
| Dissaving | 1.7 (1.4, 2.3) | <.05 | 1.8 (1.4, 2.3) | <.05 |
| Unmet social need | 1.3 (1, 1.8) | .06 | 1 (.7, 1.3) | 0.8 |
| MPI 3 Categories |  |  |  |  |
| Not multidimensionally poor/not vulnerable (Ref.) | -- | -- | -- | -- |
| Not multidimensionally poor/vulnerable | 1 (.5, 2.2) | 1 | 1.2 (.5, 2.3) | 0.8 |
| Multidimensionally poor/severely poor | 1.1 (.7, 1.7) | 0.7 | 1.1 (.7, 1.7) | 0.7 |
| Social Protections Receipt | 1.3 (.9, 1.8) | 0.6 | 1.6 (.7, 3.7) | 0.3 |
| Gender based violence | 1.8 (.9, 3.4) | 0.1 | 1.6 (.9, 2.8) | 0.1 |
| Intimate partner violence | 1.9 (1.4, 2.5) | <.05 | 1.6 (1.0, 2.4) | <.05 |

## DISCUSSION

Our findings highlight the extensive socioeconomic vulnerabilities faced by pregnant and lactating women living with HIV in Zimbabwe—challenges likely mirrored in other low-income settings in sub-Saharan Africa. Approximately half of respondents reported engaging in at least one form of dissaving. Over half of participants’ households would be considered poor or severely poor based on the multidimensional poverty index, and another 22% were considered vulnerable to poverty. More than half reported experiencing at least one form of socioeconomic vulnerability, most commonly food insecurity. These findings are consistent with the high national prevalence of food insecurity in Zimbabwe,^14^ and resemble findings across sub-Saharan Africa.

We found several associations between these measures of socioeconomic vulnerability and HIV outcomes in this population of pregnant and lactating women. In both unadjusted and adjusted models, dissaving was significantly associated with delayed presentation to antenatal care. Women who engaged in any form of dissaving had nearly twice the risk of delayed presentation. Given that dissaving may be proxy indicator for socioeconomic distress,^22^ this finding supports our hypothesis that economic vulnerability constrains engagement in antenatal care. Unexpectedly, multidimensional poverty and household size—often considered proxies for household-level deprivation—were not associated with viral load non-suppression, ARV interruption, or delayed antenatal care in either adjusted or unadjusted models. This finding may be due to our relatively small sample size.

Across multiple models, intimate partner violence (IPV) was independently associated with both HIV viral load non-suppression and delayed presentation to antenatal care, consistent with prior studies in the region. For instance, Gibbs et al. (2022) found that IPV was associated with reduced viral suppression among pregnant women in South Africa,^23^ while Aboagye et al. (2022) observed that IPV decreased the likelihood of attending a prenatal appointment within the first 12 weeks of pregnancy.^17^ These results underscore the urgent need for policies and programs to prevent IPV and support women experiencing violence, particularly those who are pregnant or lactating. Strengthening IPV screening and referral systems in antenatal and HIV care settings could improve viral suppression and maternal health outcomes.

We found that neither costs incurred with HIV care nor transportation time to clinic were not significantly associated with viral load non-suppression, ARV interruption or delayed presentation to antenatal care. It is notable that while HIV treatment in Zimbabwe is free, antenatal care and other non-HIV services often require user fees.^24^ While we had assumed that pregnant and lactating women may face additional financial barriers to care, Zimbabwe’s use of differentiated service delivery models for HIV—where clients receive 3–6 months of ARV refills per visit—may mitigate the financial strain associated with frequent clinic attendance.^25^ Our findings indicate that 40% of respondents received a three-month supply of ARV and one-third received a six-month supply. Consequently, most women only need to visit clinics twice a year, reducing transport-related barriers. Likewise, care costs were not significantly associated with HIV outcomes—possibly because ARV is free and visit frequency is low.

Despite high rates of socioeconomic vulnerability, only one in four participants reported ever receiving social protection benefits. The World Bank defines social protection as a system designed to help poor and vulnerable populations cope with crises, invest in human capital, and protect against life-course risks.^26^ Such interventions—including cash transfers, nutrition support, and job training—are key pillars of both the Joint United Nations Programme on HIV/AIDS (UNAIDS) and the Sustainable Development Goals (SDGs).^27,28^ Evidence from low-and middle-income countries suggests that social protection can reduce mother-to-child HIV transmission,^29^ improve engagement in antenatal care,^30,31^ enhance viral suppression,^32^ and decrease HIV transmission.^33^ These interventions are particularly vital in Zimbabwe, where 70% of the population lives below the Total Consumption Poverty Line and 29% lives in extreme poverty.^34^ This gap between need and coverage reflects a persistent mismatch between supply of social protections and demand, consistent with findings from other sub-Saharan African settings showing that many eligible individuals do not receive available benefits.^35^

Interestingly, we found that receipt of social protection benefits was associated with ART interruption. Although we initially hypothesized a protective effect of receiving social protection, this finding may indicate that that social protection recipients are disproportionately drawn from socioeconomically vulnerable populations already at higher risk for treatment interruption.^36^ A single, small disbursement may have limited effect, particularly if not timed to periods of acute financial hardship. Additionally, future studies should capture more granular data on the timing, frequency, and amount of social protection benefits received. Moreover, the existing landscape of social protection programs in Zimbabwe focused on educational benefits and livelihood support and may not have been designed to address the specific needs of pregnant and lactating women group.^37–39^

Our study has several limitations. All clinical outcomes were self-reported by participants, which may have introduced recall or reporting bias. The cross-sectional nature of our dataset may have reduced model sensitivity, particularly as we used generalized estimating equations (GEEs) which although suitable for clustered data, have better performance in longitudinal datasets. The wide confidence intervals in our models suggest that our sample size may have been underpowered to detect smaller effects, however, this is understandable given the formative nature of our work. Finally, because the CSS survey only captured whether a participant had ever received social protection, a critical exposure in our analysis, we were unable to assess dose–response effects or timing of benefit receipt. These limitations, we hope, can be overcome with additional research focused on addressing unmet social needs and vulnerabilities among pregnant and lactating women with HIV.

## CONCLUSIONS

This study provides novel insights into the relationship between socioeconomic vulnerability, social protection, and clinical outcomes among pregnant and lactating women living with HIV in Zimbabwe. We found that intimate partner violence was associated with HIV viral load non-suppression and delayed presentation to antenatal care, while dissaving was associated with delayed antenatal presentation. Social protection receipt, though limited in coverage, was paradoxically associated with ARV treatment interruption—highlighting the need for deeper investigation into the context and timing of such interventions.

## Appendix A

### Household size

Participants were asked how many members lived in their household, as crowding is a frequently cited measure of poverty^40,41^ which, in turn, has been shown to impact both HIV and antenatal treatment outcomes. Participants were asked how many people live in their household, and responses were categorized into the following categories: 1) 1-2 people, 2) 3-4 people, and 3) 5 or more people.

### Living with a disability

Participants were asked if they had a disability. Although the survey asks individuals to further describe their type of disability (physical, hearing, visual, mental, intellectual, or albinism), we only included whether or not individuals self-identified as having any kind of disability (yes/no).^42^

### Transportation time to clinic

Individuals were asked how long it took them to reach a health clinic from their place of residence. Responses were categorized as: 1) 30 minutes to 1 hour, 2) 1-2 hours, and 3) More than two hours. Individuals who lived within 30 minutes of the health center were grouped with individuals who did not have to travel to a health center, as this was considered relatively local in our study setting. We included this covariate because prior studies have demonstrated that distance to a healthcare center is a potential barrier to HIV^43^ and antenatal care.^44^

### Costs incurred with care

Participants were asked if they incurred costs for healthcare, either costs associated with transportation to a health care facility or fees associated with a clinic visit, as prior studies have described cost as a barrier to care.^45^ Responses were dichotomized (yes/no).

### Dissaving

Participants were asked if they or their family had experienced dissaving since March of 2020. Specifically, participants were asked if they had 1) taken out a loan, 2) sold an asset, 3) withdrawn money from savings^46^, reduced personal or household food consumption, or 5) taken a child out of school to support access to HIV care or adherence to ARVs. Measures of dissaving are frequently used in settings where it is difficult or impossible to collect data about individual or household income.^47^ Responses were dichotomized.

### Social protection intervention ever received

In the survey, participants were asked if they had ever been a recipient of a social protection intervention. Those who responded “yes” were asked to further specify which of the three main social protection interventions they had been recipients of (the Harmonized Social Cash Transfer, the Emergency Social Cash Transfer, and/or the Basic Education Assistance Model, described in further detail below). For our GEE model, we included a dichotomous variable indicating whether or not an individual had been a recipient of any social protection intervention.

### Social protection-specific program

Individuals who had been recipients of social protection interventions were asked to further specific which intervention they, or their household, had been a beneficiary of. Participants responded that they were either beneficiaries of the Harmonized Social Cash Transfer (HSCT), the Emergency Social Cash Transfer (ESCT), and/or the Basic Education Assistance Module (BEAM). HSCT is a nationally implemented cash transfer intervention targeting impoverished and food insecure households as defined by the Zimbabwean Ministry of Health and Child Care (MoHCC).^37^ HSCT is jointly funded by the government of Zimbabwe and partners, including UNICEF, and has been implemented in waves at the district level during specific time frames. ESCT was an additional cash transfer program jointly implemented by UNICEF and the MoHCC between 2020 and 2023 across three urban districts in Zimbabwe. ESCT was created in an effort to address poverty, food insecurity, and socioeconomic vulnerability that had been exacerbated by the COVID-19 pandemic, and consisted of monthly payments of $48 for eligible households.^38^ Lastly, BEAM is a government funded, nationally implemented program to help orphans and vulnerable children pay for school-related fees (e.g. tuition and exam fees).^39^ It has been primarily implemented by the Zimbabwean government.

### Unmet social needs

To evaluate unmet social needs, participants were asked if they had experienced financial or food vulnerabilities that affected their adherence to antiretroviral therapy and/or their ability to attend clinic visits. Specifically, participants were asked if they ever 1) Lacked food for themselves or for family, 2) Lacked funds for transportation to or from the health facilities, 3) Lacked funds for user fees or other costs at the health facility, or 4) Experienced any other form of socioeconomic vulnerability (described with free text). If a participant answered “yes” to any of the aforementioned questions, they would be considered to have experienced vulnerability. Responses were dichotomized accordingly.

### Multidimensional poverty

The 2022 Multidimensional Poverty Index (MPI), which was created by the Oxford Poverty and Human Development Index in partnership with the United Nations Development Program. The index uses ten indicators across three dimensions to evaluate deprivation at the household level. The three dimensions are health, education, and standards of living.^18^ Health and education each have two indicators, while standards of living has six indicators. Weight is assigned accordingly: each health indicator is weighted 1/6, while each standard of living indicator is weighted 1/18. Each of these weights is considered to be a “deprivation score,” and the sum of the indicators is the household deprivation score with a maximum deprivation score of 100%.^18^ Households with a deprivation score greater than 1/5 (20%) but less than 1/3 (33.3%) are considered to be vulnerable to multidimensional poverty. If the household deprivation score is 1/3 (33.3%) to ½ (50%), the household is considered to be multidimensionally poor. If the household deprivation score is ½ (50%) or greater, the household is considered to be in severe multidimensional poverty. For our GEE analysis, we grouped multidimensionally poor with severe multidimensional poverty to help ensure a sufficient number of participants in each category; we were therefore less likely to find associations by chance. Participants were asked about each of these ten indicators and scored according to the above described calculations.

### Intimate partner violence & gender-based violence

Participants were asked if they had experienced abuse from their intimate partner in the last three months and if they had experienced gender-based violence in the last three months. We chose to include these as covariates because current evidence suggests that both gender-based violence and intimate partner violence negatively affect HIV treatment outcomes and engagement with antenatal care.^23^ Responses were dichotomized (yes/no). Participants were also asked if they had experienced gender-based violence from anyone other than their intimate partner in the last three months, as the survey is administered quarterly. Responses were dichotomized (yes/no).

### Number of months of ARVs given

Participants were asked how many months of ARVs they were given at their last clinic visit. This was asked primarily because Zimbabwe uses a differentiated service delivery model, in which individuals receive several month’s supply of ARVs in a single clinic visit.^48^ Differentiated service delivery models have been implemented in settings where it may be difficult for individuals to access clinics on a monthly basis. Response options were 1) One month, 2) two months, 3) Three months, and 4) Six months. This covariate was only used in our descriptive statistics but was not included in the GEE analysis. The primary reason this covariate was included in descriptive statistics was to help us better describe and quantify how individuals access HIV care in our study setting.

## Data Availability

The data that support the findings of this study contain sensitive personal information and are not publicly available due to ethical and privacy restrictions. Access to the anonymized dataset may be granted to qualified researchers upon reasonable request, subject to the execution of a data sharing agreement and approval by the University of California San Francisco Institutional Review Board (IRB). Requests for data access should be directed to the corresponding author at.

## Contributors/acknowledgements

Stella Bialous, Wendy Max, Jillian Kadota and most importantly, all the participants in our surveys who dedicated their time to supporting this work. We also want to thank the ZNNP+ group who prioritizes care of individuals with HIV.

---

**Supplementary table 1.** Additional demographics and socioeconomic characteristics of pregnant and lactating women living with and receiving care for HIV in 15 districts of Zimbabwe (N = 600)

|  | N (%) |
| --- | --- |
| **Pregnant/Lactating (n = 600)** |  |
| Pregnant | 153 (25.5%) |
| Lactating | 447 (74.5%) |
| **District** |  |
| Beitbridge | 59 (9.8%) |
| Bulawayo | 13 (2.2%) |
| Bulilima | 42 (7.0%) |
| Chiredzi | 65 (10.8%) |
| Chitungwiza | 22 (3.7%) |
| Chivi | 19 (3.2%) |
| Gutu | 49 (8.2%) |
| Gwanda | 62 (10.3%) |
| Insiza | 47 (7.8%) |
| Mangwe | 26 (4.3%) |
| Masvingo | 48 (8.0%) |
| Matobo | 42 (7.0%) |
| Mwenezi | 36 (6.0%) |
| Umzingwane | 42 (7.0%) |
| Zaka | 28 (4.7%) |
| **ARV # months prescription given** |  |
| 1 month | 56 (10.4%) |
| 2 months | 9 (1.7%) |
| 3 months | 255 (47.1%) |
| 6 months | 221 (40.9%) |

**Supplementary table 2.** Demographics and socioeconomic characteristics of pregnant and lactating women living with and receiving care for HIV in 15 districts of Zimbabwe (N = 600), stratified by whether or not clients had been recipients of social protection interventions.

|  | Has never been a recipient of SP (n=481)<br>N (%) | Has been a recipient of SP (n=119)<br>N (%) | P value |
| --- | --- | --- | --- |
| **Pregnant/Lactating (n = 600)** |  |  | .9 |
| Pregnant | 123 (25.6%) | 30 (25.2%) |  |
| Lactating | 358 (74.4%) | 89 (74.8%) |  |
| **Age** |  |  |  |
| **Age Group** |  |  |  |
| Young adults | 181 | 44 | .6 |
| Middle-aged adults (20-44) | 278 | 67 |  |
| Adult (45-64) | 22 | 8 (6.7%) |  |
| **Viral Load result (within past 12 months) (n = 497)** |  |  | <.05 |
| <1000 | 257 (85.7%) | 48 (70.6%) |  |
| >= 1000 | 43 (14.3%) | 20(29.4%) |  |
| Unknown |  |  |  |
| Interruption to ARV (Yes) | 42 (8.7%) | 17 (14.3) |  |
| First Antenatal Appointment (n = 600) |  |  | .2 |
| Before 12 weeks | 354 (73.6%) | 79 (66.4%) |  |
| After 12 weeks | 127 (26.4%) | 40 (33.6%) |  |
| Household size |  |  |  |
| Household size groups |  |  |  |
| 1-2 | 88 (18.2%) | 13 (11%) | .1 |
| 3-4 | 182 (37.8%) | 44 (37%) |  |
| 5+ | 211 (43.9%) | 62 (52.1%) |  |
| Living with a disability | 11 (2.3%) | 9 (7.6%) |  |
| District |  |  |  |
| Beitbridge | 54 (11.2%) | 5 (4.2%) | <.05 |
| Bulawayo | 11 (2.3%) | 2 (1.7%) |  |
| Bulilima | 21 (4.4%) | 21 (17.6%) |  |
| Chiredzi | 53 (11%) | 12 (10.1%) |  |
| Chitungwiza | 22 (4.6%) | 0 (0%) |  |
| Chivi | 17 (3.5%) | 2 (1.7%) |  |
| Gutu | 41 (8.5%) | 8 (6.7%) |  |
| Gwanda | 50 (10.4%) | 12 (10.1%) |  |
| Insiza | 39 (8.1%) | 8 (6.7%) |  |
| Mangwe | 14 (2.9%) | 12 (10.1%) |  |
| Masvingo | 41 (2.9%) | 7 (5.9%) |  |
| Matobo | 25 (5.2%) | 17 (14.3%) |  |
| Mwenezi | 30 (6.2%) | 6 (5%) |  |
| Umzingwane | 38 (7.9%) | 4 (3.7%) |  |
| Zaka | 25 (5.2%) | 3 (2.5%) |  |
| Transportation time to clinic |  |  |  |
| 0-30 minutes | 118 (24.7%) | 25 (22%) | .9 |
| 30 min to 1 hour | 211 (45%) | 53 (46%) |  |
| 1 hour to 2 hours | 117 (24.3%) | 31 (26%) |  |
| More than 2 hours | 22 (4.7%) | 5 (4.2%) |  |
| Paid for clinic services (Yes) | 17 (3.5%) | 1 (.8%) |  |
| Paid for transportation (Yes) | 195 (40.5%) | 44 (37%) |  |
| Received ARVs at clinic visit if due (Yes) | 439 (91.3%) | 102 (85.7%) |  |
| ARV # months prescription given |  |  | .4 |
| 1 month | 50 (10.4%) | 6 (5%) |  |
| 2 months | 8 (1.7%) | 1 (.8%) |  |
| 3 months | 205 (42.6%) | 50 (42%) |  |
| 6 months | 176 (36.6%) | 45 (37.8%) |  |
| Unmet social need(Any) | 258 (53.6%) | 67 (56.3%) | .6 |
| Lack of food for participant or family | 220 (45.7%) | 63 (52.9%) |  |
| Lack of funds for transportation to or from health facility | 132 (27.4%) | 25 (21%) |  |
| Lack of funds for user fees or other costs at the health facility | 114 (23.7%) | 28 (23.5%) |  |
| Other |  |  |  |
| Dissaving (Any) | 231 (48%) | 66 (55.5%) | .1 |
| Taken out a loan | 26 (5.4%) | 4 (3.4%) |  |
| Withdrawn from personal or household savings | 86 (17.9%) | 18 (15.1%) |  |
| Sold assets | 116 (24.1%) | 38 (32%) |  |
| Reduced personal or household food consumption | 158 (32.8%) | 48 (40.3%) |  |
| Taken a child out of school | 55 (11.4%) | 16 (13.4%) |  |
| MPI 4 categories |  |  | .1 |
| Not multidimensionally poor/not vulnerable | 93 (19.3%) | 21 (17.6%) |  |
| Not poor but vulnerable | 106 (22%) | 27 (22.7%) |  |
| Multidimensionally poor | 220 (45.7%) | 46 (38.7%) |  |
| Severe multidimensionally poor | 62 (12.9%) | 25 (21%) |  |
| Social Protections Program Receipt (Ever) |  |  |  |
| HSCT (n = 119) |  | 29 (24.4%) |  |
| ESCT (n = 119) |  | 35 (29.4%) |  |
| BEAM (n = 199) |  | 57 (47.9%) |  |
| Experienced Intimate Partner Violence in past 3 months (Yes) | 40 (8.3%) | 12 (10.1%) | .5 |
| Experienced Gender-Based Violence in past 3 months (Yes) | 24 (5%) | 6 (5%) | 1 |

